# Centering healthcare workers in developing digital health interventions: usability and acceptability of a two-way texting retention intervention in a public HIV clinic in Lilongwe, Malawi

**DOI:** 10.1101/2023.01.09.23284326

**Authors:** Maryanne Mureithi, Leah Ng’aari, Beatrice Wasunna, Christine Kiruthu-Kamamia, Odala Sande, Geldert Davie Chiwaya, Jacqueline Huwa, Hannock Tweya, Krishna Jafa, Caryl Feldacker

## Abstract

**Background:** New initiates on antiretroviral therapy (ART) are at high risk of treatment discontinuation, putting their health at risk. In low-resource settings, like Malawi, appropriate digital health applications must fit into local connectivity and resource constraints. Target users’ perspectives are critical for app usability, buy-in and optimization. We describe the formative stages of the design of a two-way text-based (2wT) system of tailored reminders and adherence messages for new ART initiates and share results from key informant interviews with HCWs focused on app usability and acceptability.

**Methods:** Using a co-creation approach with clients, clinical, technical and evaluation teams and over app development, we held four informal user feedback sessions, a small pilot with 50 clients, and ten key informant (KIIs) to deepen our understanding of healthcare workers (HCWs) needs, acceptability and usability.

**Results:** Formative research informed the design of interactive client-to-HCW communication, refining of the language and timing of weekly text blast motivational messages and tailored client-specific visit reminders. Informal feedback from HCW stakeholders also informed educational materials to enhance 2wT client understanding of how to report transfers, request visit date changes and ask questions related to their visits. In KII, HCWs noted their appreciation for the co-creation process, believing that the participatory HCD process and responsive design team enabled the development of a highly acceptable and usable 2wT digital tool. HCWs also suggested future improvements to promote inclusion of clients of varying literacy levels and economic backgrounds as well as integrating with other health platforms to improve uptake of 2wT.

**Conclusions:** Inclusion of HCWs increased perceptions of app usability and acceptability among HCWs. HCWs believe that 2wT will improve on-time ART visit attendance and provide valuable early retention in care support. The co-creation approach appears successful in designing an app that will meet HCW needs and, therefore, support client adherence to visits.

**Author Summary:** People starting antiretroviral treatment (ART) are at risk of poor or non-adherence which could lead to treatment failure or drug resistance, putting their health at risk. To help improve client adherence and, therefore, client outcomes, healthcare workers and healthcare systems need user-centred digital innovations that are appropriate for low- resource settings. Using a co-creation approach between the clients, clinical, technical, and evaluation teams, we designed, developed, and optimized a two-way text-based (2wT) system of tailored reminders and motivation messages for new ART initiates at Lighthouse Trust’s public ART clinic in Lilongwe, Malawi. The application leveraged the open-source, Community Health Toolkit, and was designed based on evidence of previous 2wT success in client engagement. We describe the formative stages of app co-design and share results from key informant interviews with HCWs focused on app usability and acceptability. We detail findings on the 2wT system design, perceived strengths, weaknesses, and recommendations for improvement to inform continued optimization for scale up. Overall, the co-creation approach appears successful in designing an app that will meet HCW needs and, therefore, support client adherence to visits.

## Introduction

In sub-Saharan Africa (SSA), retention in antiretroviral therapy (ART) is an increasing challenge that threatens both individual health and attainment of UNAIDS targets to end the AIDS epidemic by 2030 [1]. With significant healthcare system constraints and formidable healthcare worker (HCW) shortages, ART services in SSA struggle to meet ambitious global UNAIDS 95-95-95 targets where 95% PLHIV know their status, 95% of those on ART and 95% of those have their viral load (VL) suppressed. In Malawi, a recent public health survey found UNAIDS target achievement at 88-87-83 [2], demonstrating a gap in attainment of critical milestones. Evidence suggests digital health interventions for direct provider to client engagement with people living with HIV (PLHIV) are highly acceptable and contribute to improved adherence to antiretroviral therapy (ART) visits [3-6], an important component of engagement in care. However, not all mHealth interventions are effective for all populations at all times [7], and even well-designed, well-researched interventions might not be effective [8-10]. Malawi’s mobile connectivity index—a composite of infrastructure, affordability, consumer readiness, content and services—was 31.6 in 2021 versus a sub-Saharan Africa regional index of 41.5, suggesting that digital health interventions may face barriers in digital innovation success [11]. Recent global guidance on app development for global health suggests that improved monitoring and evaluation (M&E) and strengthened efforts to engage diverse users in app co-design may help improve mHealth effectiveness even in low resource settings [12, 13]. A core suggestion is to increase engagement of local healthcare workers (HCWs) in app design from inception to help identify the right intervention for the specific context [14]. Adherence to those global guidelines, including emphasis on HCW user inputs, may help increase the likelihood of bringing innovations to scale [15].

Participatory human centered design (HCD) can be used to develop digital innovations that are appropriate for low resource settings and that improve client adherence to clinic visits. HCD informs digital solutions by enhancing client engagement for complex health care needs and ensuring that innovations are user-friendly for local healthcare staff [16-20]. HCD employs a specific mind- and skill-set that places users at the centre of the problem-solving process and values lived experiences to develop solutions that are meaningful to a given user context [21-22]. The co-creation process promotes gathering of feedback, analysis, and incorporation of diverse user inputs in system optimization to respond to user’s evolving needs, skills and behaviour during the product life cycle. Hallmarks of quality HCD-led digital innovations are easy to use, learnability and adaptability of the solutions to respond to changing user needs, especially for HCWs [14]. To assess those hallmarks, usability among HCWs is critical [23]. Usability includes how target innovation users employ the system in ways that demonstrate alliance between identified gaps and application of the innovation to consistently, and correctly fill those gaps [24]. Formative research and usability testing with diverse HCW users guides early, and repeated, feedback.

Lighthouse Trust (LT), the largest public provider of ART in Malawi, operates five large clinics across the country in Blantyre, Lilongwe, Mzuzu, and Zomba in collaboration with the Malawi Ministry of Health (MoH) and supports over 65,000 PLHIV in care. In Lilongwe, LT operates the Lighthouse Clinic (LH) and Martin Preuss Center (MPC), with a combined 35,000 ART clients. Both clinics employ a real-time electronic medical records system (EMRS) and implement a resource-intensive client retention program, Back to Care (B2C). B2C successfully traces clients who miss ART visits by >14 days (presumed lost to follow up [LTFU]) via phone or home visit. B2C has been successful [25-29], but gaps are growing as client volume grows. MPC, alone, has over 7,800 monthly visits: approximately 10 percent of clients are B2C-eligible per month. High B2C demand, coupled with scarce resources, results in tracing delays and ART interruptions.

Therefore, to address gaps in adherence to ART visits and reduce the B2C workload, the University of Washington’s International Training and Education Center for Health (I-TECH), Lighthouse Trust, and open-source technology partner, Medic, applied HCD to design, develop, test, and implement an innovative, appropriate, client retention system using two-way texting (2wT) between new ART clients and HCW, aiming to increase client engagement and adherence in care. 2wT builds on successful technology developed in Zimbabwe for voluntary medical male circumcision (VMMC) that found that 2wT-based follow-up between providers and clients in lieu of routinely scheduled post-operative visits was safe for clients and reduced provider workload [30]. 2wT reduced program costs [31], was usable for both clients and healthcare providers [30], and was brought to scale within routine settings [32]. The aim of the Lighthouse 2wT system is to improve early ART client retention, defined as on-time attendance for ART visits in their first year of ART care.

Using a co-creation approach between the clients, clinical, technical, and evaluation teams, we adapted 2wT for ART retention, designing a hybrid 2wT system of individualized reminders and motivational messages for new ART initiates at Lighthouse Trust’s public ART clinic in Lilongwe, Malawi. 2wT was based on the open-source Community Health Toolkit (CHT) and used widely-available SMS technology for the low resource setting. As healthcare workers (HCWs) perspectives are critical for app usability, buy-in and optimization, we sought to engage HCWs in app co-creation from 2wT conceptualization. The objective of this paper is to present the formative HCD approach and the usability assessment including feedback from HCWs to design the 2wT system for ART retention. We describe the formative stages of app co-design as informed by four groups of diverse stakeholders, a small pilot, results from key informant interviews with ten HCWs that focused on app usability and acceptability. We provide lessons learned and key successes that may inform scale-up of this or other digital health innovations to strengthen ART retention. If successful, 2wT could contribute to improving ART retention at low-cost, providing an app that could be adapted for use in routine, high-volume, public settings in Malawi or in SSA.

## Methods

### Study Setting

Lighthouse Trust is a WHO-recognized Center of excellence (COE) that operates two clinics in Lilongwe, Malawi [33]. The 2wT intervention was implemented in the largest COE, Martin Preuss Centre clinic (MPC), in Bwaila Hospital located in Lilongwe, Malawi. MPC currently has over 24,000 clients alive in care.

### Participants

The overall 2wT intervention was designed with and for new ART initiates (within 6 months of starting ART) and HCWs at MPC. HCW participants in the HCD-focused formative research and usability study were selected by purposive sampling to identify participants based on association with retention efforts, including 2wT. For the formative HCD research using informal feedback sessions, eligible HCW participants included key stakeholders such as: departmental managers, M&E team members (data officers and IT officers), Expert Clients (MPC client-to-client ART mentors), and Back to Care staff, including field tracers and retention officers. For the key informant interviews (KIIs), only HCWs actively engaged in the 2wT implementation were eligible for interviews.

### Technology Overview: The Community Health Toolkit (CHT)

2wT system was built on a foundation provided by the open-source Community Health Toolkit (CHT), a free, open-source digital health global good to advance universal health coverage [34]. The CHT Core Framework, stewarded by Medic and released under AGPL-3.0, is a global good [35] that supports community health workflows, thereby decreasing the time and resources required to build full-featured, reliable, interoperable, secure, and ready-to-scale digital health applications (apps) to improve client care rather than code from scratch. The development of the CHT adheres to WHO’s recommended principles for digital development [36]. Apps built on the CHT Core Framework are highly customizable, support multiple languages, run offline, and work with basic feature phones (via SMS), smartphones, tablets, and computers to support integrated care coordination and delivery. While the CHT is designed for community level personas, it is highly extensible through integrations with complementary Android apps, electronic medical records, messaging systems, and national health information systems to meet various needs such as bidirectional messaging, closed-loop referrals, aggregate reporting and any other additional functionality. In 2021, CHT powered applications supported 22.7 million caring activities and more than 41,400 CHWs in 16 countries across Africa and Asia [34].

### Adapting 2wT from VMMC to for ART via Human Centered Design

The core components of the target 2wT system were drawn from the VMMC intervention and adapted collaboratively by Malawian ART stakeholders and included: (1) automated workflows that send weekly motivational messages to promote wellbeing; (2) individually-tailored SMS reminders to clients with upcoming visits, with a response requested; and (3) enabling open-ended SMS texts between clients and HCWs to allow clients to reschedule visits, report transfers, or report other logistics concerns. Medic designers led participatory HCD activities, collaborating closely with a multi-disciplinary team of clients, healthcare workers, supervisors and software developers to specify the system requirements for the local context. This approach considers the motivations, concerns and behaviour of the end users and facilitators across the design process, limiting time and resources wasted on creating features that do not benefit, are not relevant, or are unacceptable to the target population [37-41]. At multiple stages, key stakeholders were presented with updated workflow drafts, including features for automatic flags for clients who miss clinic visits, task management and prioritization features, longitudinal client records, data collection forms, routine syncing, and dashboards for routine monitoring. Iterative improvements were informed by analysing users’ system usage patterns and insights gleaned during feedback sessions. Medic’s continuous HCD approach had four dynamic, non-linear phases (Figure 1):

**Fig 1.** Developing and optimizing 2wT for ART retention: Medic’s human-centered design process [16] [REDACTED]

### Data collection

Data collection took place over three distinct phases: 1) informal feedback sessions with 2wT stakeholders, 2) a small pilot, and 3) key informant interviews (KIIs).

### Phase I: formative HCD co-creation research and informal feedback sessions

We conducted four feedback sessions from October and November, 2020. Sessions included various MPC stakeholders involved in client retention to understand their needs and context, informing the 2wT design. MPC ART clients were also involved in informal feedback. 2wT HCWs conducted two practice educational sessions with 8 clients who volunteered to contribute (3 men and 5 women) to inform development and improvement of 2wT recruitment and educational materials. Documentation of formative feedback sessions was collected via notes and program reports from 2wT staff.

### Phase II: Evaluation from a small-scale pilot and subsequent 2wT app improvement

A soft 2wT pilot took place in June 2021 with 50 new ART clients, identifying strengths of the system and key areas of improvement before launch. Results during and after the pilot were discussed with 2wT HCWs, M&E staff and Medic teams via a series of five joint remote design sessions. This series of discussions focused on clients and HCWs feedback and an analysis of system usage trends. Each session built on the previous insights and inputs, focusing on optimizing 2wT for HCWs (system users, M&E and 2wT data users) and clients.

### Phase III: Usability perspectives from HCWs post-implementation

2wT launched in June, 2021. Six months later, we conducted ten KIIs with healthcare workers involved in 2wT implementation, supervision, M&E, and IT support over a period of 2 weeks using a facilitator guide. The KII guide was pre-tested and revised by the study team. A trained facilitator from Lighthouse but external to the 2wT study team conducted the interviews. Participants were recruited, briefed on the purpose of the activity and written consent obtained for recording by those who chose to participate. Each interview session took place in English and lasted for approximately 30 mins. All data collection tools and reports were uploaded to a secured folder only accessible by the study researchers.

### Data Analysis

For phases one and two, informal program reports were reviewed and synthesized by the 2wT study team. For the KIIs, we generated verbatim transcripts with Otter.ai software; recordings were deleted once transcribed and the transcripts anonymized. We conducted our analysis using NVivo 12 Pro qualitative text analysis software (QSR International. Burlington, Massachusetts). We used a few transcripts to develop and refine a codebook using inductive and deductive approaches and coded all interview scripts. The team collaboratively reviewed and discussed the resultant codes to formulate themes based on the DEPICT method of participatory qualitative analysis [42] that included HCWs in review of initial codes and preliminary results determination.

### Ethics

The parent study, including the usability and acceptability components, was reviewed and approved by the University of Washington and the Malawi National Health Sciences Research Committee. Participants in KIIs provided written informed consent. No other identifiers were produced nor utilized for formative data analysis in phases one or two.

## Results

### 2wT system flow

In brief, the 2wT system (Figure 2) is a hybrid automated and manual texting interaction between clients and HCWs that includes both weekly general motivational messages about general wellness sent to all enrolled clients and appointment reminders sent to each client in anticipation of their specific visit with a response requested. Responses yield follow up actions for HCWs such as confirming clinic transfers, visit updates and referrals (Figure 3). If a visit is missed, clients receive up to 3 additional reminders until they either return to the clinic or get referred to B2C for tracing. SMS is free for clients. Messages are sent either in English or Chichewa according to clients’ preferences, but clients can send an SMS at any time in any language. Clients can opt out of motivational and/or visit reminder messages at any time. 2wT officers usually respond within 1-2 days. Additional details on the 2wT intervention were described previously [43].

**Figure 2.**
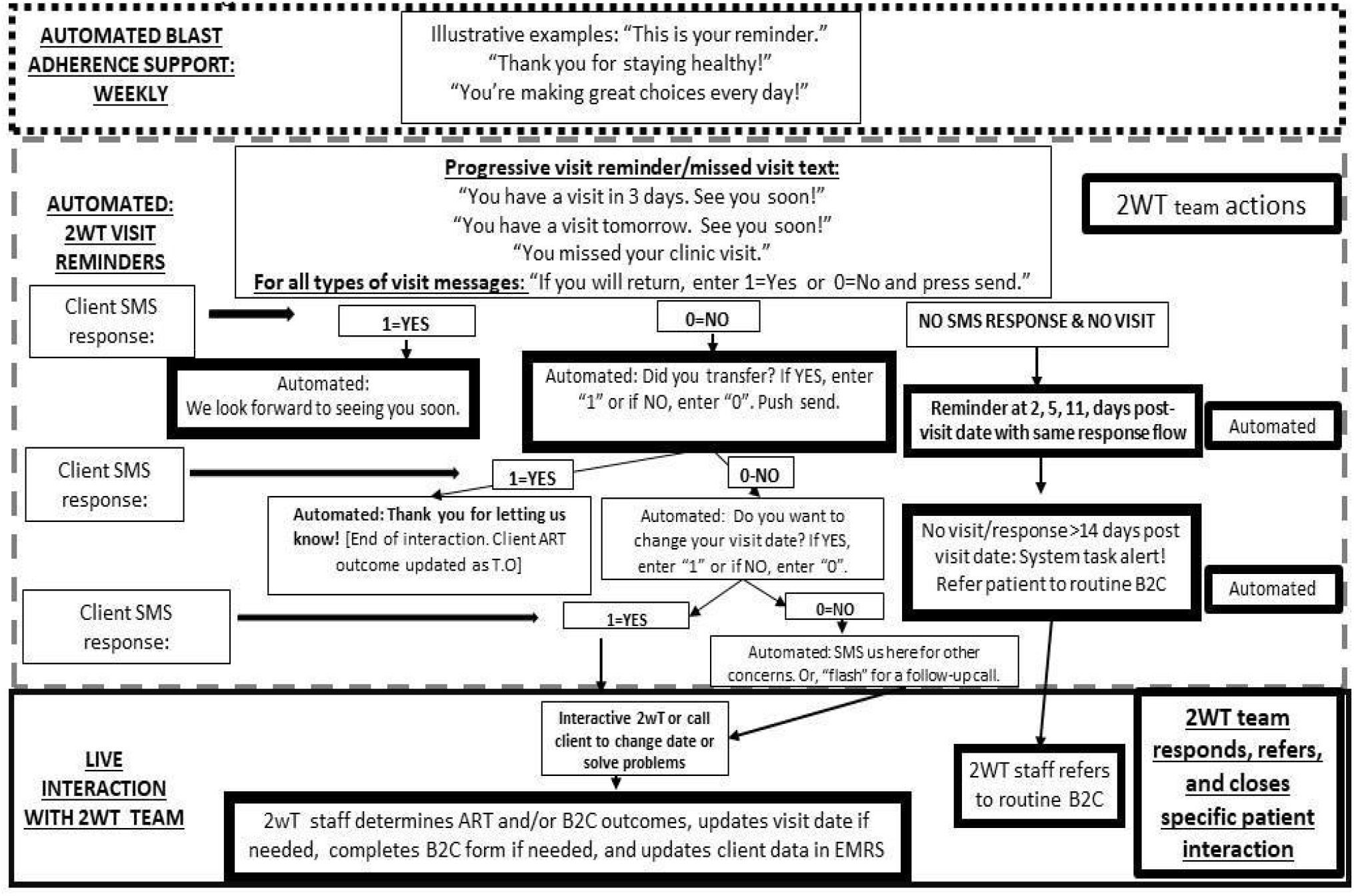
2wT flow diagram for ART retention [43]

**Figure 3.**
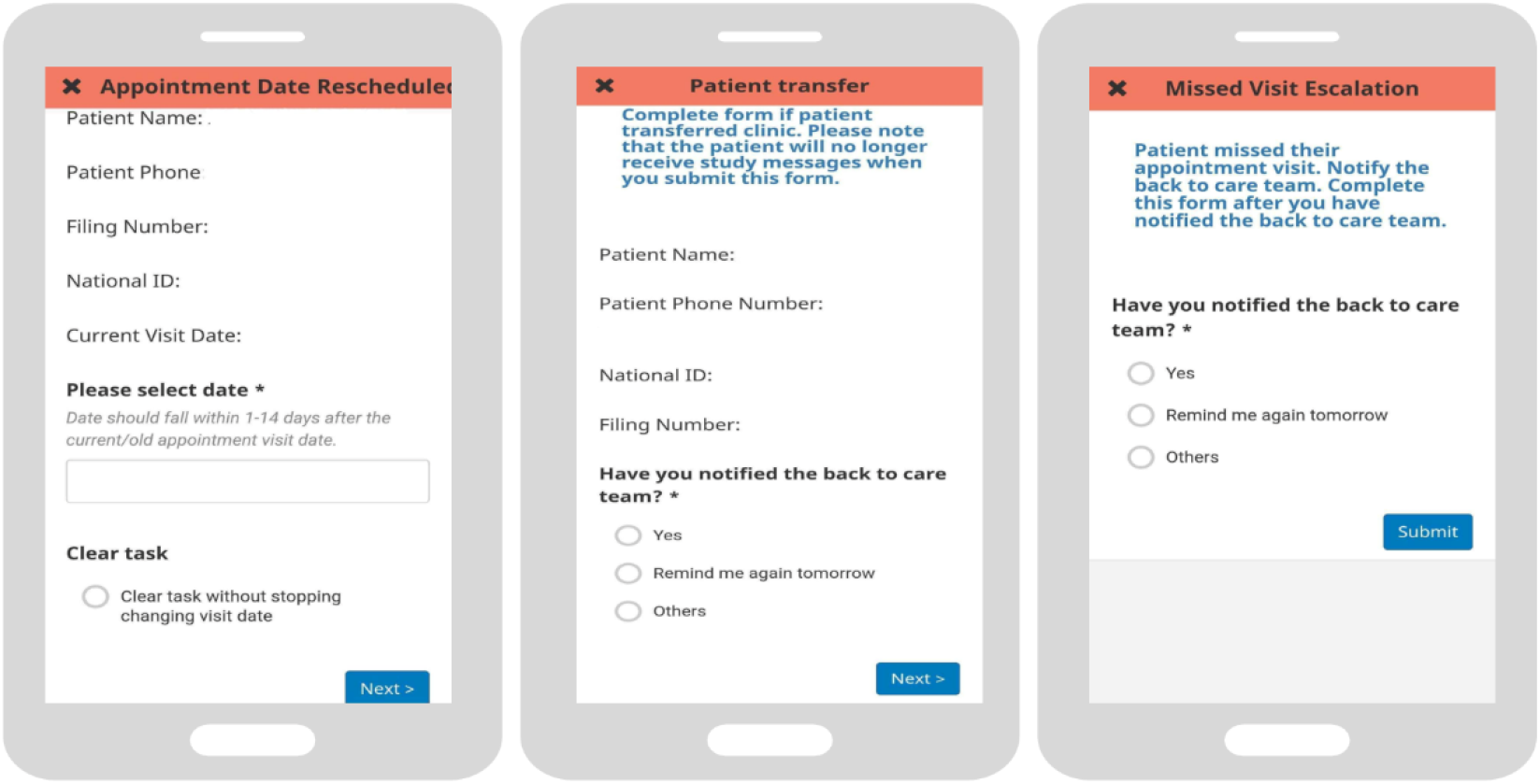
2wT actions completed by HCWs

### Phase I Outcomes: Formative HCD co-creation and informal feedback

Several key areas of client-focused improvements were identified across groups and addressed. First, for clients, 2wT is opt-in only, reducing risks of confidentiality or disclosure. To improve inclusion of, and acceptability of 2wT for clients, the level of literacy was reduced to a minimum and available in Chichewa and English, as selected by the client. Expert clients helped write, review, revise, and translate adherence and motivational messages; expert clients were identified as the best group to conduct 2wT education for new ART clients. 2wT educational materials were augmented to include a client flow chart to guide HCWs in explaining to clients how the message flow worked, how to respond, and how to stop messages. HCWs received a flip book and poster to better explain to clients how to interact with the system with illustrations for participants on how to respond with a “1” or “0” and to improve clarity on SMS communication via free text messages. The neutrality of all text messages was reviewed to ensure no reminder nor motivation words would expose the HIV status of the client (e.g., HIV, ART, adherence, clinic, etc.). With the opt-in and clear education for clients before voluntary enrolment, valid concerns were reduced about others/or spouse seeing the text messages where there is risk of disclosure or abuse. HCWs also informed the SMS timing for clients who were potential defaulters (those who missed their visits by >14 days), landing on SMS reminders at 3 times after a missed visit and before 14 days.

For HCWs, additional sensitization confirmed that 2wT would augment, not replace, other early retention interventions. To reduce redundancy between 2wT-focused interventions and other expert client-led or phone reminder systems, SMS reminder timing would be aligned with other B2C interventions for standardization. Client recruitment would take place during routine B2C recruitment and coincide with routine locator and tracing form completion, reducing impact on Lighthouse routine operations. 2wT activity flows and HCW workload were discussed, including discussions of potential workload increases (2wT interaction on the system) and decreases (reduced tracing if clients were maintained in care). As a result, one HCW was designated as the “2wT officer,” and accountable for daily review and responding to client SMS in the morning and afternoon, following up with calls when needed. A set of standard operating procedures (SOPs) for 2wT to EMRs data exchange were created, providing steps to update visit dates, de-identify shared data, increase security, reduce system errors and smooth data flow. Concerns about reminding clients accidentally after they actually attended a visit were diminished due to regular synchronization of data in EMR and 2WT system to prevent or reduce false referral for missed visit reminders or tracing.

### Phase II Outcomes: Pilot evaluation and subsequent 2wT app improvement

The pilot revealed challenges in the EMRs to 2wT data exchange that were addressed via joint working sessions focused on data exchange using automated and manual approaches. THe resulting process was a one-way flow of data from the EMR to 2wT allowing only offline EMR data access for 2wT. The team explored the data pipelines and information needs, including how records would be identified and uniquely mapped to the specific clients files. Protections for client data were strengthened, including data export for consenting participants with limited variables relevant to appointment dates and ART outcomes only. Concerns around dealing with EMR outages led to the inclusion of data collection forms which were used to manually update appointment dates. Outcomes captured in 2wT were retrospectively entered into the EMR system. For cost reduction and efficiency, the study team applied for a short code using both Airtel and TNM cellular services for optimization, a decision that allowed all clients to communicate for free via 2wT but delayed the launch by several months. Additional concerns about potential delays or failed SMS resulted in several additional M&E processes including measuring the timeliness of SMS interactions, assessment workload, and message failure rate reviews. System usage analysis of messaging interactions revealed errors in the system such as clients continuing to receive messages after they had transferred clinics and restrictive SMS broadcasting rules that return messages as expired or failed if a phone number is absent on the network or off for over 12 hours.

### Phase III Outcomes: Usability perspectives from HCWs post-implementation

#### Advantages of the 2wT platform: client-focused

Almost all health care workers (HCWs) noted that the automated client visit reminders helped clients remember their visits. KII5 reported that, *“the feedback that we’re getting from participants is that previously they could forget the day they were supposed to come to the clinic*.*”* HCWs also believed that reminder messages encouraged adherence to medications, by helping them plan ahead to make time and transport arrangements:

> *“They have good reminders, they are preparing on time, they might be having a problem of transport but because of the reminders it is giving them time to prepare that in the next three days they should look for transport so that they can come to the clinic in time*.*” [KII5]*

HCWs approved of the weekly motivational SMS with non-HIV related health education or uplifting messages, reporting that the feedback they received from clients was positive. “*Apart from being reminded that this is your appointment date there are some other bonuses like they also receive motivational texts. So, they also learn something in this. So, they are killing with two or more birds with one stone*.” [KII6]. Another put the support expressed by clients succinctly: *“The motivational messages also makes them feel that they are not alone, that we are together with them in every season of life”* [KII3].

Several HCWs expressed that clients may prefer SMS reminders over phone call reminders as it offers privacy and allows clients to read and respond to messages in their own free time.

> *“The back to care team, they used to call before the 2wT system, maybe three or four times, they would call the participant to come for their visit. So, during those frequent calls to the participants, at times, they [*clients*] were not at a good place to pick the call*… *with the* [2wT] *intervention it is easier. It’s because as I mentioned earlier, that even if they call the participants several times, but that participant maybe in a group, is in a bus, cannot respond to those calls, but with the intervention, the messages, they just respond right away [KII5]*.

#### Advantages of 2wT platform: HCW-focused

For HCWs, themselves, almost all believed that 2wT would reduce the workload in several ways. First, HCWs largely noted the expectation that if 2wT was successful fewer clients would be referred to Back to Care for missed visits and active tracing:

> *“One can already tell the benefit is to say they will reduce our workload in terms of the number of clients that we must chase up to them whether through phone or through field tracing. So that’s one of the major benefits that probably say apart from producing other costs as well. As we are following up clients there are costs that are incurred for us to trace them so that will also probably be reduced*.*”[KII7]*

Also, 2wT provides a list of clients who missed visits by 14 days without having to run a manual query of the EMRs for clients who miss visits. One B2C staff member noted, “*If those clients fail to come, these guys, the 2wT guys, immediately give us [*Back to Care] *those names. So, we traced those clients before they become defaulters” [KII8]*. 2wT efficiency gains could reduce the time needed to begin tracing since Back to Care would be “*notified much faster about a missed visit than waiting to extract the file for them after maybe three weeks or four weeks*” [KII2}.

Another KII specified how identifying outcomes proactively via 2wT could reduce work that would typically be completed by active tracing:

> *And we even get some outcomes from them through the SMS interactions (someone had died, some are transferred out) from the responses that are given to the 2wT officer. So we had some outcomes, ART outcomes, even before we initiate the tracing*.. *we just transferred those outcomes into the tracing forms*.” [KII 8]

#### Challenges of 2wT: client-focused

HCWs also considered challenges from the perspective of clients. For example, “*some of them also change numbers, which affect the study because it seem as if the clients are not responding to the messages* [KII4].” Although 2wT is opt-in (clients ask to participate), another HCW stated that some clients still may not want to come to the clinic: “*They have read it. They have received and read it, but they just ignored it*” [KII7]. Another explained the depth of the potential problem:

> *“In villages, they [2wT clients] may keep their phones off for some time. I think the challenge would be on the technology side, where if we do not have a good percentage of people who do not own their own phones, and if they own their own phones, and if they keep it on all the times…Therefore, that has to be explored. Like the behavior in terms of people, the cultural behavior of people having their phones on, people owning their own phones, people changing their phone number*.*” [KII1]*

Several HCWs mentioned client confidentiality concerns, reducing uptake, noting that some clients worry that messages from the aggregator (short code/sender number) may be recognized or deduced from others, potentially revealing that a client is on treatment. One HCW, reflecting on why some clients may not opt into 2wT offered that:

> *“Some think that it is not safe in terms of confidentiality, especially when they are at home, even though we explain to them the way the text messages were designed. They still feel like it is not a good thing, maybe there are many people who have access to their phone. Some people think a lot and may have questions as to what is going on (if they see the messages). There cannot be more that 10 people in that category*.*”[KII3]*

HCWs were disappointed by how many clients were unable to participate in 2wT for failing to meet eligibility criteria including having a phone and basic literacy. One HCW noted their surprise at the number of clients without phones, stating that, “*on the ground, with the data have, we find that most people they don’t have phones. So, it means those people that might not benefit from this system*.*”[KII4]* HCWs also expressed that the program’s inclusion criteria locked out clients who would benefit from the intervention, for example, “*some of them do not read or write. They are willing but they are they are failing to be enrolled*.*”[KII6]* Even literate clients may need quality 2wT education at enrollment to help them understand how to interpret, and successfully respond to, the message prompts.

#### Challenges of 2wT: HCW-focused

Although discussion of 2wT strengths intimated widespread support for the intervention, HCWs discussed several key challenges. First, although 2wT may reduce the early retention effort, several HCWs expressed concern that there is potential for increased responsibility and workload to manage the system and interact with clients.

> *My concerns for expanding study, it will need staff as I said. And also workload will be high. We will have time to check the texts because we will be receiving a lot of texts on daily basis. So they need somebody to be there fully, to reply the texts. In short it will consume time and the human resource. [KII6]*

Health workers complained about technological challenges, some internal to the system and some due to broader network challenges. Early on, in the first weeks, there were concerns that the system was not sending messages on time, so someone needed to “*check if the messages are able to pass through the systems…the delivery of the messages to the client*.*”* [KII1]. In addition to concerns about the aggregator working correctly, there is the challenge of timely and complete syncing of Lighthouse to 2wT data. If Lighthouse experiences an issue with the EMR like a power outage that causes visits to not be entered on time or there are delays between EMR to CHT syncing, “*some participants end up receiving a missed visit escalation, a missed visit reminder yet they reported*.*” [KII2]*. Many noted that improved integration with the EMRs would help solve several challenges including smoothing exchange between 2wT and EMRs on visit attendance, easing updating of tracing outcomes, and automating updates between systems rather than manual file transfer.

### Health care workers suggestions on 2wT scale-up

Several HCWs noted the need for additional staff if the 2wT intervention expands to include all clients and not new initiates, recognizing that, “*if we are to expand, we will be looking at the whole cohort which is too big, which means everyone has to pass through this [2wT enrolment] room making it to be a lot of work*.*”[KII9]*

For 2wT system improvements, HCWs suggested that 2wT should enrol multiple numbers per client, including numbers from different telecoms.

> *“On the message, you’ll find they have used another number, on the registration they use another number, when they are coming, they also use another number. Yeah, so we need to identify that this is the very same person but how are we going to identify that? It’s by asking them when they come for enrolment. Do you have other numbers, then we can be able to know*.*” [KII5]*.

There were also policy related suggestions. Several hoped that clients without phones could enroll for 2wT via phones of friends or buddies by allowing clients to enroll in reminders via a trusted number:

> *We would meet some clients who said that “it is my husband who has a phone” or “I live with my mother”, so if it was possible that we could be using phones of those that they feel they could disclose to, this can make it easy to reach a lot of contacts because those who do not have phones have a guardian with a phone*.*” [KII10]*

To improve 2wT acceptability for clients, and to engage more clients who could benefit from 2wT, there will need to be outreach and advocacy to make sure clients know about 2wT. *I think when it is being scaled up; there will be that sensitization to make people aware of the programme, both for the clients, and for the providers, that they know what is going on” [KII1]*. 2wT messages could be further simplified so that very low literacy clients could enrol. One HCW explained:

> *My observation is that not everyone can get involved in [2wT], just like another study, not everyone is eligible. But 2wT is restricted to a certain category of people that have the capability of having a phone…Considering our literacy rate in Malawi, and also the economic challenges that we are facing, some clients won’t be able to get into 2wT. Though they would love to be part of the study, but they wouldn’t qualify* [be eligible] [KII7]

Other interesting comments outside of key themes from HCWs are worthy of consideration. First, although SMS may be less expensive when compared to the time and costs incurred making phone calls or travelling to trace clients in the community, one HCW expressed that what *“has not been really explored more is on the SMS. The cost of SMS for Malawi, they are quite a bit higher as compared to other countries” [KII1]*. Another HCW observed that 2wT might reduce opportunities for in-person counselling since when a client “*missed an appointment, he/she is supposed to be counselled by the back to care team. Now with texting that means the physical interaction will not be there, so no kind of counselling will be provided to the client for future appointment*.*” [KII7]*.

Lastly, HCWs noted that clients may want more information from the 2wT than simply to discuss visits. *“Some clients may have [ART] questions that need to be responded to through the 2wT system, but we do not want to discuss clinical things using the system because the personnel in the two-way texting and reply to the texts are not clinical personnel*.”[KII9]

## Discussion

The 2wT app reflected the realities of scarce HCW and financial resources and appears both usable and acceptable for HCWs in this routine ART clinic setting in urban Malawi. HCWs believed that 2wT was safe, easy to use, low cost and valuable to them. The use of highly participatory, iterative HCD approaches throughout the planning, co-creation, prototyping, pilot, and implementation phases helped ensure that app design aligned with HCW stakeholders and matured according to World Health Organization guidance [44]. By observing users in their environment, working together to define the problems to be solved, collaborating with users to generate and test ideas, and evaluating the appropriateness of the prototypes with users, HCD helped closely align the evidence-base to user priorities and context [45]. All three phases of the HCW-focused formative work provided insights into 2wT strengths and weaknesses that inform optimization and scale of 2wT and similar ART retention app in the SSA region.

First, 2wT reflected local HCW preferences and priorities in accordance with the principles of digital development [36], likely aiding 2wT buy-in. HCWs and stakeholders recognized the need for intervention to improve client retention without increasing their workload, making the 2wT approach highly useful. HCWs also appreciated that the app could help clients report transfers or delay visits in ways that were accessible and appropriate for

Lighthouse client users [46] while reducing unnecessary tracing and streamlining the generation of the defaulters list – two recognized burdens. Multiple rounds of feedback with HCWs and clients reflected best practices in mHealth design and helped tailor 2wT flow to healthcare user preferences [47]. HCWs also liked the automated support for clients to remember and plan for their upcoming appointment visits, hence reducing the potential needs for other retention efforts. Lastly, HCWs supported the SMS approach for its privacy and convenience as clients could read and respond to SMS at any time as compared to phone calls.

Second, usability and acceptability assessment provided HCWs with an opportunity to identify several 2wT challenges. First, although 2wT will likely help prevent clients from being traced and reduce wasted tracing efforts, at scale 2wT could still increase workload as other digital technologies studies reported [19,48-49]. Responding to clients via SMS interactions, timely identification of more defaulters, and more clients identified for tracing could create need for additional support staff to help 2wT expand. Additional M&E of the 2wT system will further determine cost impact and help quantify the workload impact. Second, HCWs were concerned about client eligibility and access as 2wT required ownership or primary use of a mobile phone and enough literacy to use SMS. Subsequent reduction of message complexity, allowing of shared phones, and enrolling all interested clients (not just new initiates) were suggested for future. A robust client education system is also needed to confirm that clients with low literacy levels understand the system prompts and are empowered to reach out to HCWs when need arises. Lastly, technology still poses challenges for digital health. There is a need for integrating standalone digital health ecosystems to ensure seamless data flow [50]. Lack of 2wT integration with EMRs was highlighted as a pain point for HCWs.

Several suggestions could help bring 2wT more smoothly to scale. 2wT is intended to complement, and not replace, other ongoing retention activities. Once established as effective (forthcoming), 2wT should be included as part of comprehensive retention efforts, allowing clients to choose a retention support system (2wT, phone calls or expert client home visits) that meets their needs. Not all clients restrict their SMS to visit related information; developing a triage and referral for clinical questions from 2wT clients to the call center is worthy of consideration. On the technology side, dashboard development should streamline client tracking while completion of the interoperability pipeline between 2wT and the EMRs would smooth daily updates to client visits and ART status. Additional layers of artificial intelligence or natural language processing advances could also support imputation of next visit dates, reduce workload, and efficiently triage for follow-up.

## Limitations of this study

The formative study focussed primarily on HCW experiences and not clients. Clients are the focus of separate usability testing, forthcoming. Formative work in intervention development is inherently biased towards creating momentum for continued iterations and improvements; however, the authors and analysts worked to create balance in reporting both success and weaknesses of the 2wT approach. The effectiveness of 2wT of client retention is still unknown, reducing the impact of the usability and acceptability testing. Despite these limitations, emphasis on the role of formative research and centering of HCWs in HCD is an important contribution to continued strengthening of locally-relevant and -optimized digital health innovations.

## Conclusion

HCWs have specific needs and valuable visions of how digital health tools could aid in easing their workload, demonstrating the value of HCD for digital health interventions. Engaging HCWs throughout the design process promotes ownership and acceptability, informing software that responds to the specific needs and challenges of the contexts. The adaptation of the 2wT platform for direct provider to client engagements from short term (voluntary medical male circumcision, 14 days follow up) to longer term (ART) presents an opportunity to consolidate lessons learned and build a stronger, more flexible 2wT system that can strengthen direct communication pathways for longitudinal care. The success of 2wT for improved retention could lead to adaptation for other health care contexts that could benefit from improved interactions between HCWs and clients along the continuum of care.

## Data Availability

Complete transcripts contain data that is sensitive or includes identifying information. The edited transcripts will be available on a case by case basis after reviewing all materials for any potentially identifying details. Interested researchers may contact the corresponding author, CF, at for copies of the transcripts. Or, researchers could also contact Jane Edelson, Regulatory Specialist at the UW, for access to the transcripts. Data described in the manuscript, code book, and analytic code will be made available upon request and completion of a relevant data-sharing agreement.

## Acknowledgements

The authors wish to acknowledge the contributions of the 2wT team: Femi Oni, Adnan Alhassan, Mourice Barasa, Edwin Kagereki, Raymond Mugwanya, Susan Maigua, Antony Khaemba, Isaac Odala, Joseph Chintedza, Aubrey Kudzala, Dumisani Ndhlovu and Daniel Mwakanema, MPC retention staff, MoH collaborators and participants.

## Contributions

All authors contributed to the writing of the manuscript, provided significant inputs into all the drafts of the manuscript, approved this version to be published and agreed to be accountable for all aspects of the work.

## Supporting information

S1 Figure 1. Developing and optimizing 2wT for ART retention: Medic’s human-centered design process [16] [REDACTED]

**S2 Figure 2.**
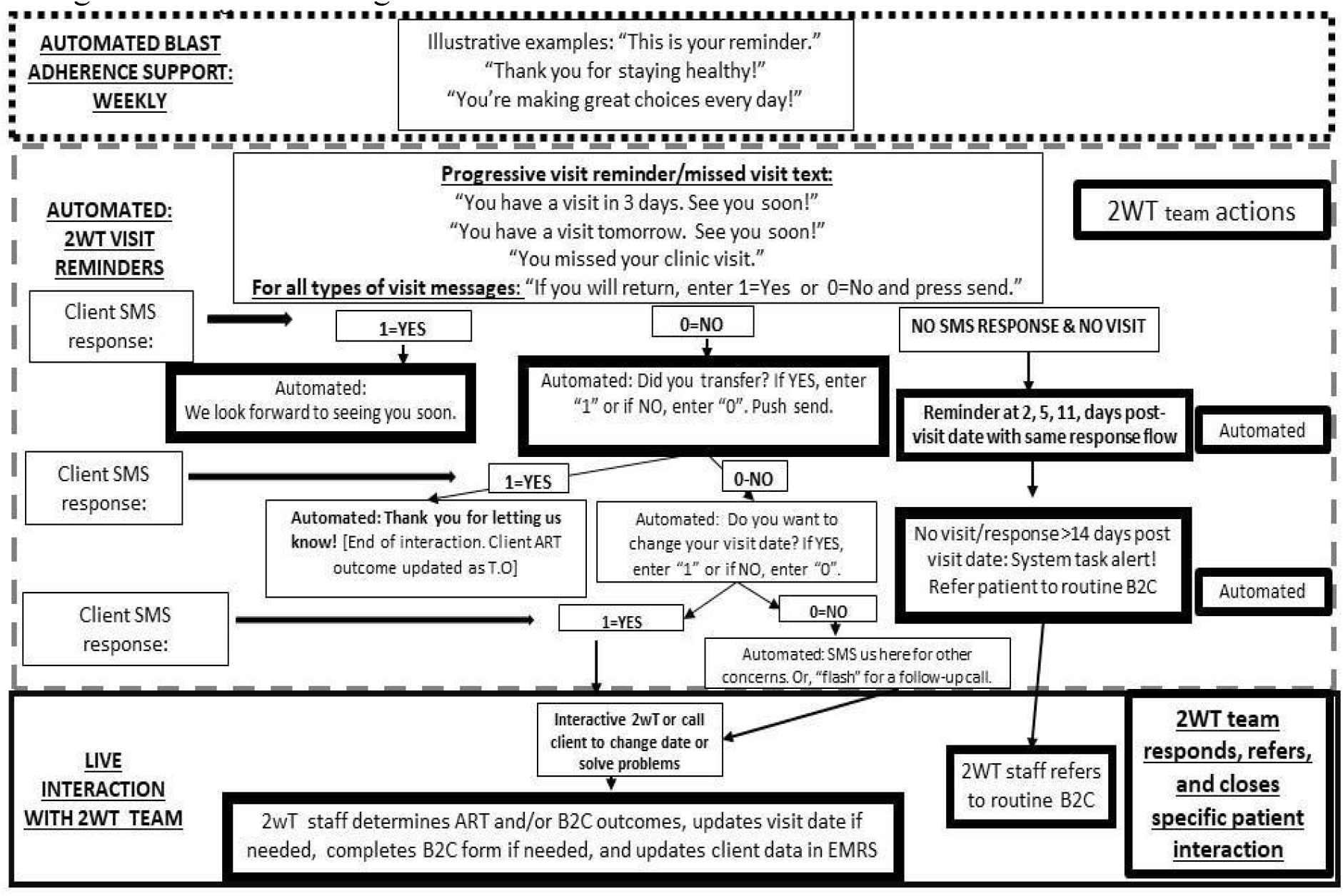
2wT flow diagram for ART retention

**S3 Figure 3.**
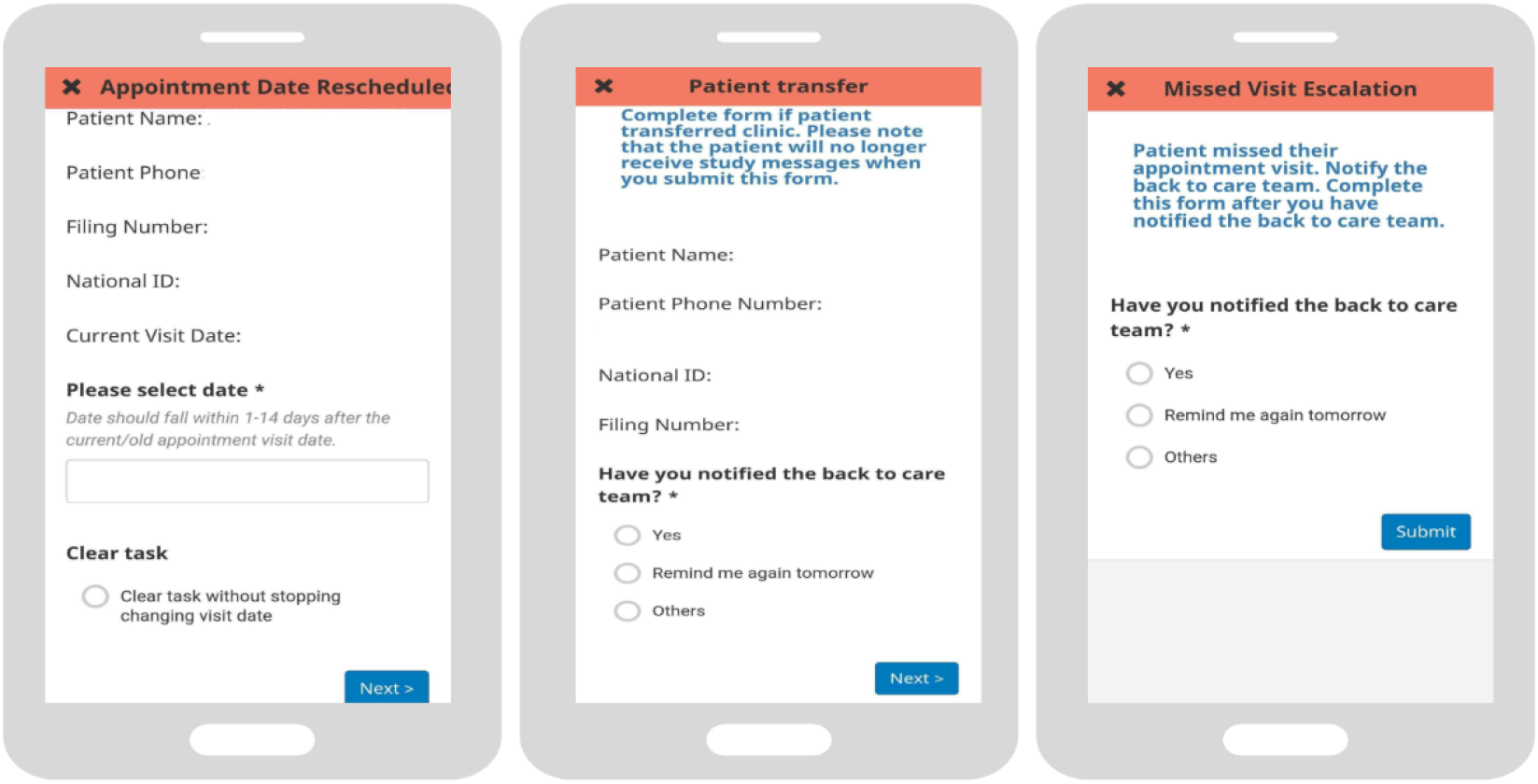
2wT actions completed by HCWs

**S1 Table.** Emergent themes.

| **THEME** | **DEFINITION** |
| --- | --- |
| **MAJOR** |  |
| Potential reduced workload | Reduced workload for the B2C team |
| Adherence benefits for clients | Support for clients to maintain adherence |
| Privacy issues for clients | Clients feel someone can see the messages and figure it out regarding ART |
| 2wT excludes some clients | Some clients who would benefit from 2wT are ineligible to enrol |
| Technology issues | System or network connectivity issues affecting the 2wT platform or electronic medical records system (EMRS) |
| Strengthen 2wT education for clients | Strengthen client understanding of the 2wT messages and usage |
| **MINOR** |  |
| Address 2wT interoperability/tech concerns | Address 2wT technical concerns regarding 2wT interoperability with the EMRs |
| Review cost considerations | Review cost considerations of the SMS platform |
| Supportive motivational messaging for clients | Motivational messaging strengthened client- health provider relationship |
| Diversify messaging to include healthy behaviors | Include other healthy living behaviors that support adherence |

